# The Effectiveness of Web-based Self-Compassion Training on Caregiving Burden and Quality of Life of Family Caregivers of Patients with Advanced Cancer: A Study Protocol for a Single-blind, Two-arm Parallel Randomized Controlled Trial

**DOI:** 10.64898/2026.10.27.26366594

**Authors:** Anita Pouzesh, Zahra Alizadeh, Mansoureh Ashghali Farahani, Naimeh Seyedfatemi, Masoud Roudbari, Mohammadreza Khanmohammadi, Mojtaba Habibi Asgarabad

**Author notes:** **Correspondence:** Zahra Alizadeh, Nursing and Midwifery Care Research Center, Health Management Research Institute, School of Nursing and Midwifery, Iran University of Medical Sciences, Tehran, Iran.,; Mojtaba Habibi Asgarabad, Department of Psychology, Norwegian University of Science and Technology, Trondheim, Norway.

## Abstract

Family caregivers of cancer patients, especially advanced cancer patients, face a significant load of care and also decline in life quality. Caregiving challenges are related to feelings of guilt, self-criticism, and shame. Self-compassion is also linked to reduced negative emotions and increased effective coping strategies. Due to time constraints and limited access to in-person support, web-based training can provide an accessible and cost-effective option for caregivers. This single-blind randomized controlled trial evaluates whether web-based self-compassion training based on Compassion-Focused Therapy improves quality of life and reduces the burden of care among Family Caregivers of patients with advanced cancer. A total of 96 caregivers aged 18 and above in oncology and palliative care units will be randomly assigned to the Compassion-Focused Therapy (CFT) group or the Treatment as Usual (TAU) group. The CFT group will receive eight sessions of web-based self-compassion training over four weeks (two sessions per week), while the TAU group will receive usual care. Data will be collected using the Self-Compassion Scale (SCS), the Caregiver Burden Inventory (CBI) to assess caregiving burden and the Caregiver Quality of Life Index–Cancer Scale (CQOLC) to assess life quality in three phases: baseline, immediately after completion of the training and four weeks after completion of the training. In addition, two researcher-developed instruments will be used: The Compassion-Focused Therapy Satisfaction Questionnaire and the Compassion-Focused Therapy Adherence Scale. Data will be analysed using linear mixed-effects modelling in R to evaluate the effectiveness of the CFT treatment. This study may assess whether web-based self-compassion training leads to a reduction in caregiving burden and an improvement quality of life for family caregivers of patients with advanced cancer.

## Introduction

Based on statistical data, after cardiovascular disease, cancer is the most life-threatening disease in the world [1]. In Asia, for example, among individuals aged 0-85 years, incident cancer cases are estimated to be around 16,162,830 between 2022 and 2045 [2, 3]. In Iran, incident cancer cases in 2024 were 156,463 [4], and this number is predicted to increase to about 210,701 in 2027 [5]. Cancer diagnosis, disease management, and treatment are extremely stressful for patients that constantly expose them to various types of financial and non-financial pressures [6-8]. This disease affects both patients and their family caregivers [8].

Family caregivers are essential in managing the physical and psychological symptoms associated with cancer [9]. Family caregivers include spouses, relatives, or friends who devote significant time (months or years) to providing health care and take on physically, socially, emotionally or financially demanding activities [10, 11]. Caring for patients presents a variety of challenges for caregivers [12]. Including: career and financial challenges [13], physical fatigue [14], decreased quality of life [15], psycho-emotional conflicts [16], Pressure to gain social acceptance [17], mental fatigue [18], lack of social support [19], social isolation [20], feeling ignored [21], stress process [22], cancer taboo [23], and heavy caregiving burden [24]. Caring for a terminally ill patient is one of the most difficult aspects of providing care [20]. The relatives of patients suffer from psychological distress, emotional problems and caregiving burden throughout the course of the disease [25].

Caregiving burden is a sophisticated and multidimensional meaning that includes negative emotions experienced by caregivers due to the social, economic, psychological, and physical, aspects of the care process [26]. Patient-related factors such as: male gender [27], patient functional status [28], poor physical and mental health [29], number of comorbidities [30], confusion [31], degree of dependency [30], and younger age [27] and caregiver-related factors such as: lower education [32], duration of care [30], longer hours of care [32] and limited support [32] both affect the severity of caregiving burden. Caregiving responsibilities have negative impacts on the mental health and quality of life of caregivers. There is a negative association between burden of care and quality of life [33]. As the burden of care increases, the life quality decreases significantly [34].

Quality of life consists of physical and psychosocial dimensions [35]. Caregiving burden affects both the quality of life of caregivers and their patients [36]. Because it reduces the quality of care that caregivers provide to patients and affects the recovery process of patient from both physical and psychological aspects [37]. Therefore, implementing targeted interventions to reduce the caregiving burden and enhance the quality of life of family caregivers seems to be very necessary.

According to the findings of the studies, interventions such as caregiver education and psychosocial support are necessary to reduce caregiving burden, because psychosocial factors have a significant impact on the severity of caregiving burden [38]. Also, the implementation of psychoeducational interventions is suggested as a useful and cost-effective strategy for reducing the caregiving burden and enhancing the quality of life of family caregivers. [8]. Cognitive-behavioral therapy [39], acceptance and commitment therapy [40] and self-compassion training [41] are among the psychological interventions that have been utilized to reduce the burden of caregiving for caregivers of patients with cancer. Among them, self-compassion training has been examined in numerous studies (Table 1).

According to a study, self-compassion training for caregivers of older adults with dementia has been associated with reduced caregiving burden and increased use of effective coping strategies to manage caregiving stress [42]. Furthermore, a study conducted by Neff et al. (2015) showed that parents of children with autism who possess higher levels of self-compassion reported lower levels of stress and depression, as well as greater life satisfaction and hope. [43].

Self-compassion means being supportive and kind to ourselves when we experience pain or suffering. It is an effective way to deal with distressing thoughts and feelings, which ultimately associated with mental and physical health [54]. Self-compassion is a form of communication with oneself in difficult situations that includes three main components: 1-being aware of when we are in distress and responding in a balanced way to difficult experiences; 2-understanding common humanity in difficult moments and recognizing that we are not alone in our suffering; and 3-the ability to be kind and understanding to ourselves in difficult moments [55]. Two main self-compassion training have been developed so far:1. Mindful Self-Compassion training [56] and 2. Compassion-Focused Therapy [57]. Gilbert, the theorist behind Compassion-Focused Therapy [58], believes that CFT is effective, particularly for individuals with severe self-criticism and shame by activating the soothing and safety system (rather than the threat system) [57]. Compared with cognitive behavioral therapy, compassion-focused therapy considers positive emotions to be more important during the therapeutic process and emphasizes improving the way individuals relate to their psychological problems [59]. The findings of three studies have shown that increased self-compassion is linked to reduced feelings of guilt, self-criticism and depressive symptoms, and promotes psychological well-being [60-62]. Self-compassion may be a better therapeutic approach than traditional cognitive therapies for self-critical individuals because it activates their soothing system more effectively [63].

Family caregivers of cancer patients have reported that they often feel guilt and self-blame due to an inability to provide sufficient, high-quality care to the patients and to support them [64]. These caregivers often neglect their own personal and health needs and even feel guilty when taking care of themselves or enjoying personal moments [44, 65]. Research has shown that caregiver guilt is positively correlated with caregiver burden; therefore, a dyadic "patient-caregiver" assessment approach may reduce caregiver guilt and alleviate caregiver burden [66]. Self-compassion can reduce the intensity of guilt by cultivating a more compassionate relationship with yourself [67]. Therefore, self-compassion training may be effective in decreasing caregiving burden and increasing their quality of life by reducing these negative emotions.

Self-compassion training can be delivered to caregivers in person or online. Using digital platforms to develop health interventions has several advantages over traditional methods, including maintaining participant anonymity, wider accessibility, and reduced cost and time [68]. Web-based interventions are very suitable and valuable due to time flexibility and easy access when face-to-face meetings are not possible [69]. According to findings of two systematic reviews, psychoeducational interventions delivered via videoconferencing can overcome barriers to attending in-person sessions and help improve caregiving skills, self-care, and the well-being of informal caregivers [70]. Furthermore, digital health interventions can address some limitations of traditional face-to-face interactions and enhance caregivers’ access to self-care services [71].

However, several studies in Iran have examined the effectiveness of self-compassion training on caregivers or parents of cancer patients in person [41, 52, 72, 73]; few studies have investigated self-compassion training as an internet-based intervention for cancer patient caregivers. Although research on internet-based interventions for cancer caregivers is expanding, the impact of these interventions on "caregiver burden" and "quality of life" remains unclear. [74].

Therefore, the development of comprehensive and tailored web-based interventions to reduce caregiving burden and improve the quality of life of caregivers of cancer patients in Iran seems essential. The main goal of this study protocol is aimed at investigating the effectiveness of web-based self-compassion training on the caregiving burden and quality of life of family caregivers of patients with advanced cancer.

The hypotheses of this study state that following an 8-session CFT program, caregivers in the CFT group will report:

A. more progress in self-compassion than those in the TAU group as assessed by the Self-Compassion Scale (SCS)
B. greater reduction in caregiving burden than those in the TAU group as assessed by the Caregiver Burden Inventory (CBI).
C. greater increase in quality of life than those in the TAU group as assessed by the Caregiver Quality of Life Index–Cancer Scale (CQOLC).

## Method

### Design

This single-blind randomized controlled trial study with two-arm and a parallel-group design compares a CFT program with a TAU group. The design of the research was in line with the SPIRIT 2025 guidelines [75]. At the beginning demographic data and also assessments at the beginning (T1), immediately after completion of the training (T2) and 4 weeks after completing the training (T3) will be collected. Figure 1 presents the study timeline (The schedule of enrolment, interventions, and assessments. Note: CFT: Compassion-Focused Therapy; SCS: Self-Compassion Scale; CQOLC: Caregiver Quality of Life Index–Cancer Scale; CBI: Caregiver Burden Inventory; CFT-SQ: Compassion-Focused Therapy Satisfaction Questionnaire; CFT-AS: Compassion-Focused Therapy Adherence Scale) while Figure 2 illustrates the CONSORT flow diagram. This study has an IRCT registration under the identifier IRCT20260203068753N1.

**Figure 1.**
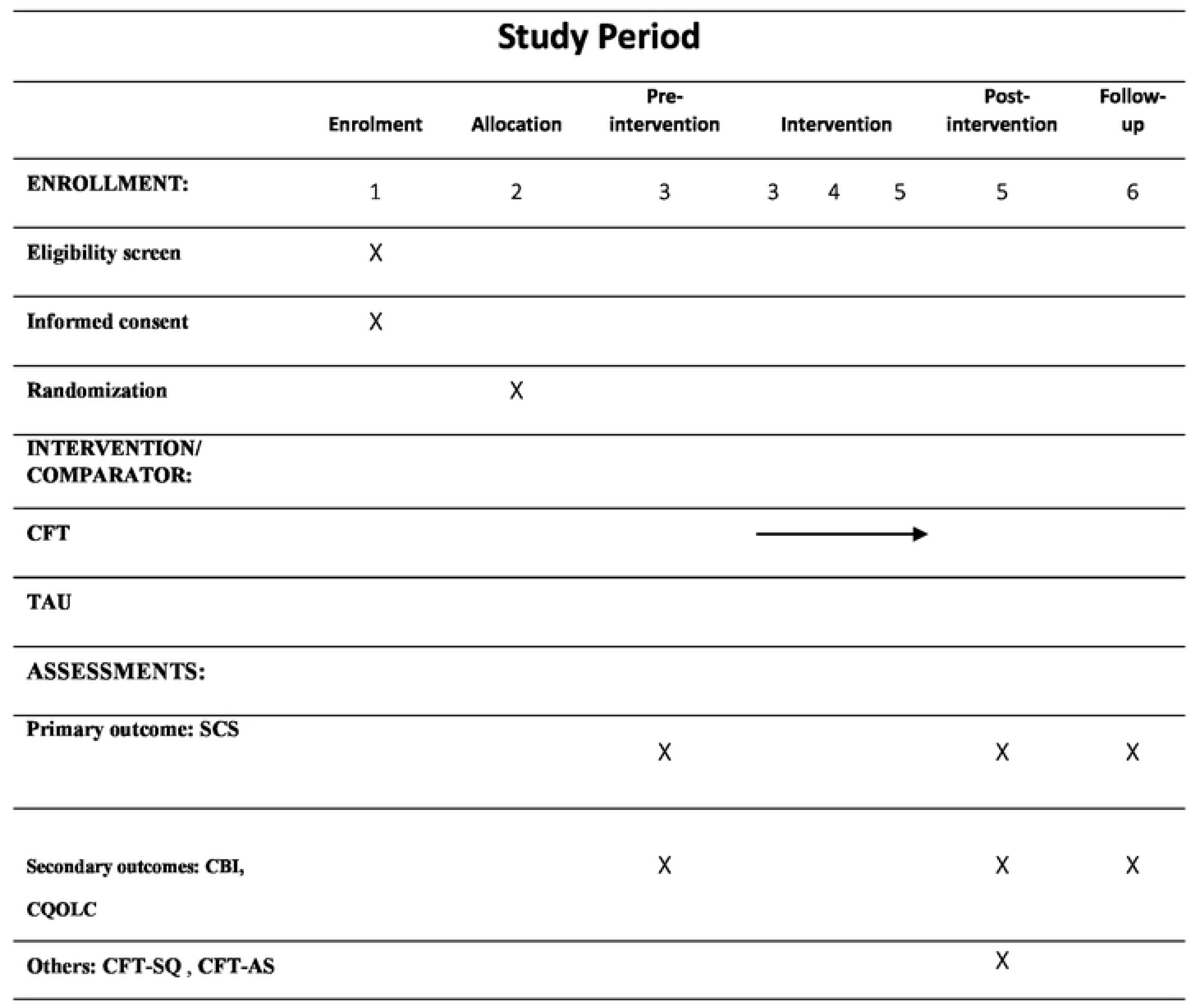
The schedule of enrolment, interventions, and assessments. Note: CFT: Compassion-Focused Therapy, SCS: Self-Compassion Scale; CQOLC: Caregiver Quality of Life Index-Cancer Scale; CBI: Caregiver Burden Inventory; CFT-SQ: Compassion-Focused Therapy Satisfaction Questionnaire; CFT-AS: Compassion-Focused Therapy Adherence Scale)

**Figure 2.**
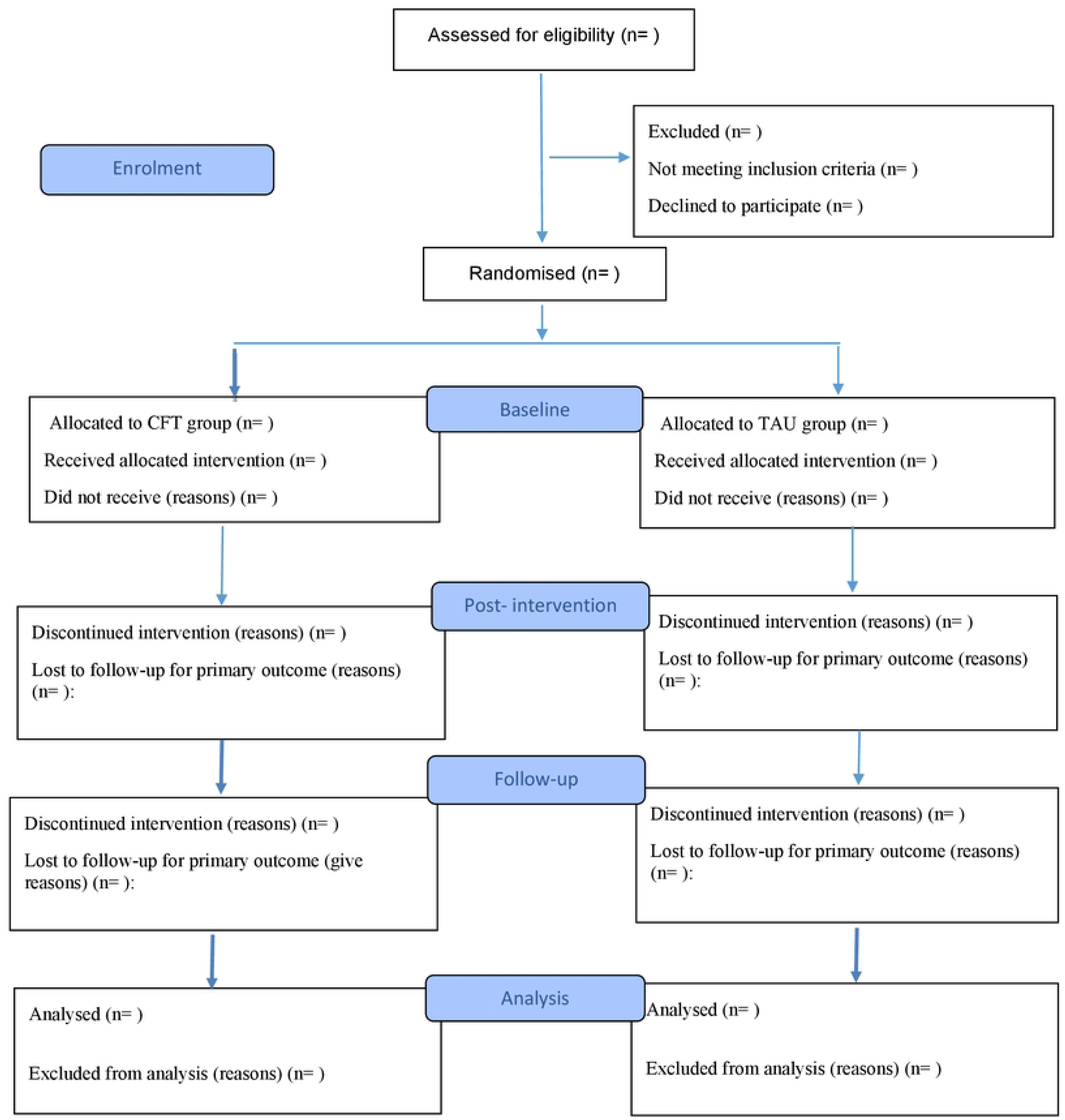
Consort Diagram of Study Protocol.

### Sample Size

We used G*Power to determine the sample size for this study [76], To perform this calculation, the following parameters were considered: effect size (f) = 0. 5, type I error (α) = .05, power of the test= 0.80, degree of freedom (df) = 2. Initially, we estimated the required sample size to be 80 for 2 groups. Considering the possibility of sample attrition during the study and in order to maintain adequate statistical power, 16 more people will participate in the study; therefore, the final sample size will be 96 people.

### Participants, Recruitment, and Randomization Procedures

The study population will consist of caregivers whose patients are admitted to the oncology and palliative care wards of two hospitals affiliated with the Iran University of Medical Sciences (Rasoul-e Akram and Firouzgar hospitals). In this study, patients with advanced cancer will be identified based on the diagnosis of the treating oncologist. After identifying eligible patients, their family caregivers will be assessed for inclusion and exclusion criteria. Inclusion criteria for caregivers included being 18 years of age or older of each gender [44], providing direct care to patients with stage III or IV cancer for at least six months [78], acting as the primary caregiver of a patient with cancer [47], being able to read and write [49], being able to communicate verbally and complete questionnaires, having a smartphone [47], being able to use a smartphone, having access to stable internet and virtual platforms [79], being able to complete educational assignments, having no history of psychiatric disorders[80], and not taking psychiatric medications when the study was being done (according to caregiver self-report). Exclusion criteria will also include death of the patient with cancer, receiving other psychological treatment during the study or within the 6 months prior to enrollment, failure to complete more than three of the seven recommended tasks, and the caregiver’s desire to withdraw from the study. If family caregivers meet the inclusion criteria, the research team will provide them with detailed information about its objectives, methodology and beneficial and adverse effects. Written informed consent is essential for all caregivers before enrollment and the collection of baseline data.

After completing the baseline assessments, caregivers will be randomized equally to the CFT or TAU group (1:1 ratio) using a four-way block randomization sequence generated by R software by an independent statistician. An independent researcher who has no involvement in participant recruitment or outcome assessment will ensure allocation concealment by keeping the randomization sequence and group allocation hidden from other members of the research team until the initial assessments are completed. After randomization, the data analyst will be the only person to remain blinded to group allocation, because caregivers and researchers cannot be blinded. Unblinding will occur only if required for participant safety or requested by the Research Ethics Committee. All instances of unblinding will be documented.

### Feasibility and accessibility Measures

To assess the feasibility of the CFT program, the rate attendance at virtual sessions, the rate of caregiver attrition, and the completion of assignments will be calculated by the researcher. To assess the acceptability of the CFT program, caregivers’ satisfaction will be measured using an item at the end of the study (How helpful was the CFT program?) responses will be scored on a 5-point Likert scale from 1 = (completely helpful) to 5 = (completely unhelpful).

### Procedures

At baseline, family caregivers will complete demographic, Self-Compassion Scale (SCS), Caregiver Burden Inventory (CBI), and Caregiver Quality of Life Index–Cancer (CQOLC) questionnaires. These questionnaires will take approximately 15 to 20 minutes to complete.

In order to implement the intervention, a web-based educational system will be designed so that participants can access the educational content through different tools, like computers, tablets and cellphones. The usability, efficiency and technical performance of the system will be evaluated and approved by relevant experts before the study. Caregivers in the intervention group will receive the web-based self-compassion training via this platform. The educational content will include narrated educational videos, audio files (podcasts), and structured educational modules. The duration of each educational video will be approximately 15 to 20 minutes and each audio file will be a maximum of 15 minutes. The intervention program will consist of eight educational sessions, which will be delivered over four weeks, two sessions in a week. For each session, practical exercises and assignments appropriate to the educational content will be provided to participants to provide the opportunity to apply the learned skills in real caregiving situations.

In order to facilitate communication with participants and resolve potential technical issues, a communication group will be created within one of the internal messaging software platforms. In addition, the researcher will be in regular contact with participants via phone calls and online communication throughout the intervention period to answer questions and potential issues, as well as to monitor their level of participation and adherence to the training program.

TAU group participants will receive only usual care during the study period and will not be provided with any educational intervention related to the study. Outcomes will be assessed using online questionnaires at two time points: immediately after completing of the training and four weeks after completion of the training. In addition, two researcher-developed instruments: The Compassion-Focused Therapy Satisfaction Questionnaire to assess participants’ satisfaction with the intervention and the Compassion-Focused Therapy Adherence Scale to assess the researcher’s adherence to the training program’s content will be used. In this study, usual care was selected as the comparator because it reflects current clinical practice for family caregivers of patients with advanced cancer and enables assessment of the incremental effect of the self-compassion education intervention. Caregivers and the general public were not involved in the planning of this study.

### TAU group

According to the routine care protocols of both hospitals, all caregivers of patients with advanced cancer will receive patient care training, which include: information about the disease and its progression, symptom assessment and management, proper medication administration, daily patient care, nutritional care, effective communication with the patient, management of common caregiving challenges, and how to contact and communicate with the healthcare team. Caregivers who wish to receive psychological supports may consult the hospital-based psychologist. Caregivers in this group will receive only patient care training and will not undergo psychological interventions during the study period.

### Compassion-Focused Therapy (CFT)

The self-compassion training intervention in this study will be based on the Compassion-Focused therapy approach designed by Paul Gilbert [81]. This approach aims to strengthen self-compassion, reduce self-criticism and shame, and improve emotional regulation ability, providing individuals with a set of training and exercises. This training program will be designed and implemented in the form of eight sessions, each session having specific educational objectives and practical exercises tailored to improving self-compassion skills.

In the initial session, caregivers will be introduced to the general structure of the program, the basic concepts of compassion and self-compassion, types of compassion, and how important is it to cultivate self-compassion in everyday life. The second session is dedicated to introducing the characteristics of compassionate people, and the personal and psychological benefits of self-compassion.

In the third session, topics such as the multiple mind, the old and new brain, the evolutionary functions of the brain, the concept of the “tricky brain,” and the difference between the threat-oriented mind and the compassion-oriented mind will be discussed. The goal of this session is to increase the awareness of participants about the origins of their emotional and behavioral reactions. In the fourth session, participants will be introduced to the three emotion regulation systems: the threat system, the arousal system, and the safety and comfort system. It will also be examined which of these systems are activated in difficult and stressful caregiving situations and the safety and comfort system can be strengthened and activated through self-compassion exercises.

Sessions five through seven focus on teaching and practicing practical self-compassion skills. In these sessions, techniques such as mindfulness, being compassionate to others and ourselves, compassionate reasoning, the multiple selves technique, Rhythmic Relaxation Breathing, three-chair technique, compassionate attention, compassionate perception, compassionate imagery, compassionate feeling, compassionate behavior, compassionate letter writing are introduced and practiced practically. Finally, in the eighth session, techniques for applying self-compassion as a routine in daily life and performing activities throughout the day with a focus on self-compassion will be introduced. A summary of the content of the training sessions is presented in Table 2.

The content validity of the self-compassion training program will be reviewed and approved by 10 Iran University of Medical Sciences experts and faculty members, including supervisors and referees, faculty members of the psychiatry nursing department, clinical psychologist, oncologist, and specialists in the field of cancer and palliative care.

### Protocol Adherence

To ensure intervention fidelity, first, all study personnel will receive standardized training before participant enrolment and the educational intervention will be implemented by a researcher who has undergone the necessary specialized training in the field of self-compassion. Second, the researcher’s adherence to the intervention protocol during the implementation of the intervention will be assessed by supervising experts using the researcher-developed Compassion-Focused Therapy Adherence Scale. Corrective feedback will be provided if any deviations from the protocol are observed. Third, all produced educational content, including narrated educational videos, podcasts and structured educational modules will be reviewed and approved by experts before use. Any necessary revisions will be made based on the feedback provided.

## Measures

### Demographics

Demographic information of caregivers will be gathered through a self-report questionnaire. This questionnaire has been developed based on a review of reliable scientific sources, including relevant research articles in the field of study. The demographic variables examined include age, gender, marital status, education level, employment status, duration of direct care of the patient, and family relationship or relationship of the caregiver to the patient. The aforementioned data will be collected at the baseline assessment stage (T1).

### Primary Outcome Measure

**Self-Compassion Scale (SCS)** developed by Neff in 2003 [82], has 26 items with 6 subscales: Self-Kindness with 5items, Self-Judgment with 5 items, Common Humanity with 4 items, Isolation with 4 items, Mindfulness with 4 items, Over-identification with 4 items. Each question is scored on a 5-point Likert scale from 1 (almost never) to 5 (almost always). The Cronbach’s alpha in the original scale ranged from 0.75 to 0.81 and 0.92 for the total scale. The scale showed high test–retest reliability (0.93). In the Iranian version, Cronbach’s alpha for the total scale reported as 0.78 and for subscales ranging from 0.79 to 0.93 [83].

### Secondary Outcome Measures

**Caregiver Burden Inventory (CBI)**, developed by Novak and Guest in 1989 [84] is a 24 items that consists of five subscales: developmental burden, time-related burden, social burden, emotional burden and physical burden. All subscales contain 5 items except physical burden which consists of four items. These items are scored on a 5-point Likert scale from 0 (never) to 4 (almost always). Subscale scores range from 0 to 20 for developmental, emotional, time-related, and social burden, and from 0 to 16 for physical burden. Higher scores indicate more caregiver burden. The Cronbach’s alpha for the original version of the CBI has been reported to be acceptable ranging from 0.69 to 0.87 and 0.80 for the total scale. In the Iranian population, the Persian version showed excellent reliability, 0.93 for total Cronbach’s alpha and 0.70 to 0.91 for subscales. At a two-week interval, test-retest reliability has also been reported to be acceptable (ICC = 0.96) [85].

**Caregiver Quality of Life Index-Cancer (CQOLC)** design by Weitzner et al. [86] which is a 35-item instrument with multiple dimensions for cancer caregivers to measure their quality of life, including 10 items for caregiving burden, 7 items for disruption of daily living, 7 items for positive adjustment, 3 items for financial concerns and 8 items for other psychological and physical factors such as sleep disturbance. Each question is scored using a 5-point Likert scale ranging from 0 (not at all) to 4 (very much). The total score ranges from 0 to 140. The original version of the CQOLC demonstrated good psychometric properties, with a Cronbach’s alpha coefficient of 0.90 and a test–retest reliability coefficient of 0.95 [87]. The Persian version of the CQOLC that was translated and validated by Khanzari et al. [88] includes four subscales: emotional burden subscale with 14 items, lifestyle disruption subscale with 9 items, positive adjustment subscale with 8 items and financial concerns subscale with 3 items. An additional item (35) assesses the family’s perceived interest in caregiving and is included in the total score but not assigned to any subscale. The internal consistency of the Persian version has been reported to be satisfactory, 0.89 for the total Cronbach’s alpha and 0.72 to 0.90 for the subscales.

**Compassion-Focused Therapy Satisfaction Questionnaire (CFT-SQ)** is a researcher-developed questionnaire that will be used to assess participants’ satisfaction with the educational content. This questionnaire is adapted from the Mindfulness Program Satisfaction Questionnaire (MPSQ), which is itself derived from the 8-item Client Satisfaction Questionnaire [89]. It consists of eight questions that scored on a 4-point Likert scale (1 to 4) and the total scores range from 8 to 32. According to Matsubara et al. [90], the Cronbach’s alpha for both the Cebuano and Waray versions of the CSQ-8 was higher than 0.80, which is acceptable.

**Compassion-Focused Therapy Adherence Scale(CFT-AS)** as a researcher-developed scale based on the MBCT-C Adherence Scale will be used to assess the researcher’s adherence to the training program’s content. MBCT-C-AS is an unpublished scale developed by Semple and Sears [91]. This researcher-developed scale consists of 20 questions, each scored on a 3-point Likert scale ranging from 0 =Not at all to 2=Clear. Higher scores represent more adherence by the researcher. The researcher-developed scale evaluates the researcher’s adherence to the core principals of teaching self-compassion, as well as the structure and sequence of the sessions.

### Ethical Considerations

This study has an IRCT registration under the identifier IRCT20260203068753N1. The ethical principles of Helsinki (1964) [92] and its subsequent revisions, particularly the 2008 revision form the basis for conducting this study [93]. Written informed consent will be required for all participants prior to the enrolment. Individuals will participate entirely voluntarily and may leave the study at any time, for any reason and without facing any adverse consequences or impact on the usual care they receive. To ensure participant confidentiality and privacy, all gathered data will be coded, securely stored, and used solely for research purposes. No one will have access to the study data except the researchers. Following completion of the study and data collection, individuals in the control group will have access to the educational content of the web-based Compassion-Focused Therapy. Any amendments to the study protocol will require approval from the principal investigator and the Research Ethics Committee of Iran University of Medical Sciences before the corresponding changes are made to the trial registration in the Iranian Registry of Clinical Trials (IRCT).

Ethical approval for this study was obtained from the Research Ethics Committee of the School of Nursing and Midwifery at Iran University of Medical Sciences (approval No. IR.IUMS.NURSE.REC.1404.021). Trial conduct, protocol adherence, participant recruitment, and data quality will be monitored by the main investigator under the supervision of the Research Ethics Committee of Iran University of Medical Sciences.

### Statistical Strategy

First, we will screen the data for completeness, accuracy of entry, and coding; we will then analyze the data distribution and identify potential outliers using descriptive statistics and visualizations such as histograms and box plots. Statistical model assumptions will also be checked prior to the main analysis. Valid data points will not be excluded solely on the basis of unusual scores. We will analyse the data using linear mixed-effects modelling in R. Data will be imported from SPSS into R after data collection is completed. Participants will be coded according to study allocation (Compassion-Focused Therapy [CFT] vs. Treatment as Usual [TAU]). Outcome variables will be converted from wide to long format to represent the three assessment time points: baseline (T1), right after the intervention (T2), and one-month follow-up (T3).

Separate linear mixed-effects models will be fitted for each outcome variable, including self-compassion, caregiver burden, and caregiver quality of life. In these models, study group, assessment time, and the group × time interaction will be specified as fixed effects, and to account for the correlation between repeated measurements for each individual, a random intercept will be included for each participant. The main focus of the analysis will be the group × time interaction, which will assess whether changes in outcome measures over time differ between the CFT and TAU groups.

Models will be estimated using the maximum likelihood method with the lmerTest package in R. Type III tests for fixed effects will be used to evaluate whether outcome scores are different between two groups, changes over time, and show different patterns of change between two groups. We will use partial eta-squared (η²p) or standardized regression estimates to report effect sizes, where appropriate. Estimated marginal means will be calculated using the emmeans package. Post hoc comparisons will examine between-group differences at each assessment point, within-group changes across assessment times, and between-group differences in change from before intervention to immediately after completion of the training, before intervention to 4 weeks after completion of the training, and immediately after completing of the training to 4 weeks after completing of the training. Holm-adjusted p-values will be used to control for multiple comparisons.

Participants will be analyzed based on their initial assigned group, in accordance with the intention-to-treat (ITT) principle. Furthermore, mixed-effects models will be used to handle missing outcome data, assuming that the data are missing at random. We will perform Sensitivity analyses, such as per-protocol analysis, to evaluate the robustness of the results and variables such as age, caregiving burden score, and quality of life score will be reported as mean and standard deviation, while variables such as gender, marital status, and education level will be reported as frequency and percentage. No interim analyses or stopping guidelines are planned.

## Result

the initial screening of participants has not yet begun, and the necessary technical facilities and requirements for implementing the intervention are being prepared. After the recruitment and screening process of participants is completed, the implementation of the intervention will begin.

## Discussion

As cancer progresses, patients become increasingly dependent on the support of others [14]. Among them, family caregivers play a vital and essential role in providing care [94]. This heavy responsibility is often associated with a significant caregiving burden, reduced quality of life [95, 96] and frequent experiences of feelings such as shame, guilt, and self-blame in caregivers [64, 97]. Therefore, quality of life and caregiving burden are two important targets for supportive interventions.

Self-compassion, as an important psychological mechanism, activates the safety and comfort system [57] and is effective in reducing shame [60], guilt [98], and self-criticism [99]. CFT, which was designed for individuals experiencing shame and self-criticism [57], provides a suitable framework for this intervention. Delivering this intervention in a web-based format offers significant advantages over traditional face-to-face methods, including greater efficiency, time flexibility, lower cost, wider access, and the possibility of training in discrete time intervals [100].

This study has important methodological strengths, including the design of a randomized clinical trial, focusing on a vulnerable population of family caregivers, using a digital approach to increase accessibility, and planning for a one-month follow-up. Also, we will use standardized instruments to measure burden of care and life quality, which will strengthen the validity of the results. However, the implementation of this study faces three major limitations. First, there is the possibility of self-report bias due to the use of questionnaire instruments. To reduce this limitation, the presence of the researcher in the initial briefing sessions, providing clear instructions, emphasizing the confidentiality of information, and selecting valid and reliable instruments have been considered. Second, virtual education may reduce the motivation and commitment of caregivers to perform home exercises. To address this issue, regular telephone follow-ups, sending daily or weekly reminders via SMS, and creating a virtual group to share experiences, challenges, and successes are planned to strengthen peer and researcher support. Third, the need for stable and adequate-speed internet access. This limitation will be managed by considering stable internet access as one of the study entry criteria and providing the necessary technical training to address common problems in the virtual group.

Finally, this study aims to fill the gap in providing evidence-based interventions for family caregivers of these cancer patients in Iran.

### Clinical Implication

If web-based Compassion-Focused Therapy proves effective, the consequences of this study could provide preliminary evidence supporting the use of this approach in nursing support interventions and palliative care services. Furthermore, the findings could help in designing and developing digital support programs aimed at reducing burden of care and improving the life quality for these caregivers.

## Protocol version

Protocol identifier: CFT-Caregiver Trial, Protocol version: 1.0, Date: October 2026

## Data sharing

Not applicable

## Ethics Approval

This study has received ethical approval from the Ethics Committee of Iran University of Medical Sciences (Ethics Code: **IR.IUMS.NURSE.REC.1404.021**). Written informed consent will be obtained from all participants prior to enrollment.

## Data availability statement

No datasets were generated or analyzed during the current study. All relevant data from this study will be made available upon study completion.

## Dissemination policy

The results of this study will be disseminated through publication in peer-reviewed journals and updating the clinical trial registry. Participants may request a summary of the study findings after study completion.

## Acknowledgements

The authors express gratitude to all hospitals that agreed to assist in this study. The authors assume full responsibility for the content.

## Trial Registration

The present study was registered in the Iranian Registry of Clinical Trials on 2026-02-09 **(IRCT)** (code: **IRCT20260203068753N1).**

## Contributions

**Conceptualization:** Anita Pouzesh, Zahra Alizadeh, Mojtaba Habibi Asgarabad.

**Formal analysis:** Zahra Alizadeh, Masoud Roudbari

**Investigation:** Anita Pouzesh

**Methodology:** Mansoureh Ashghali Farahani, Naimeh Seyedfatemi, Mojtaba Habibi Asgarabad.

**Project administration:** Anita Pouzesh, Zahra Alizadeh.

**Resources:** Anita Pouzesh, Mojtaba Habibi Asgarabad, Zahra Alizadeh.

**Software:** Zahra Alizadeh, Masoud Roudbari

**Supervision:** Zahra Alizadeh, Mojtaba Habibi Asgarabad,

**Validation:** Anita Pouzesh, Zahra Alizadeh.

**Visualization:** Anita Pouzesh, Zahra Alizadeh.

**Writing – original draft:** Anita Pouzesh, Zahra Alizadeh, Mansoureh Ashghali Farahani, Naimeh Seyedfatemi, Mohammadreza Khanmohammadi, Mojtaba Habibi Asgarabad, Masoud Roudbari

**Writing – review & editing:** Anita Pouzesh, Zahra Alizadeh, Mansoureh Ashghali Farahani, Naimeh Seyedfatemi, Mohammadreza Khanmohammadi, Mojtaba Habibi Asgarabad, Masoud Roudbari

All authors read and approved the final manuscript.

## Appendix 1

**Table 1.**
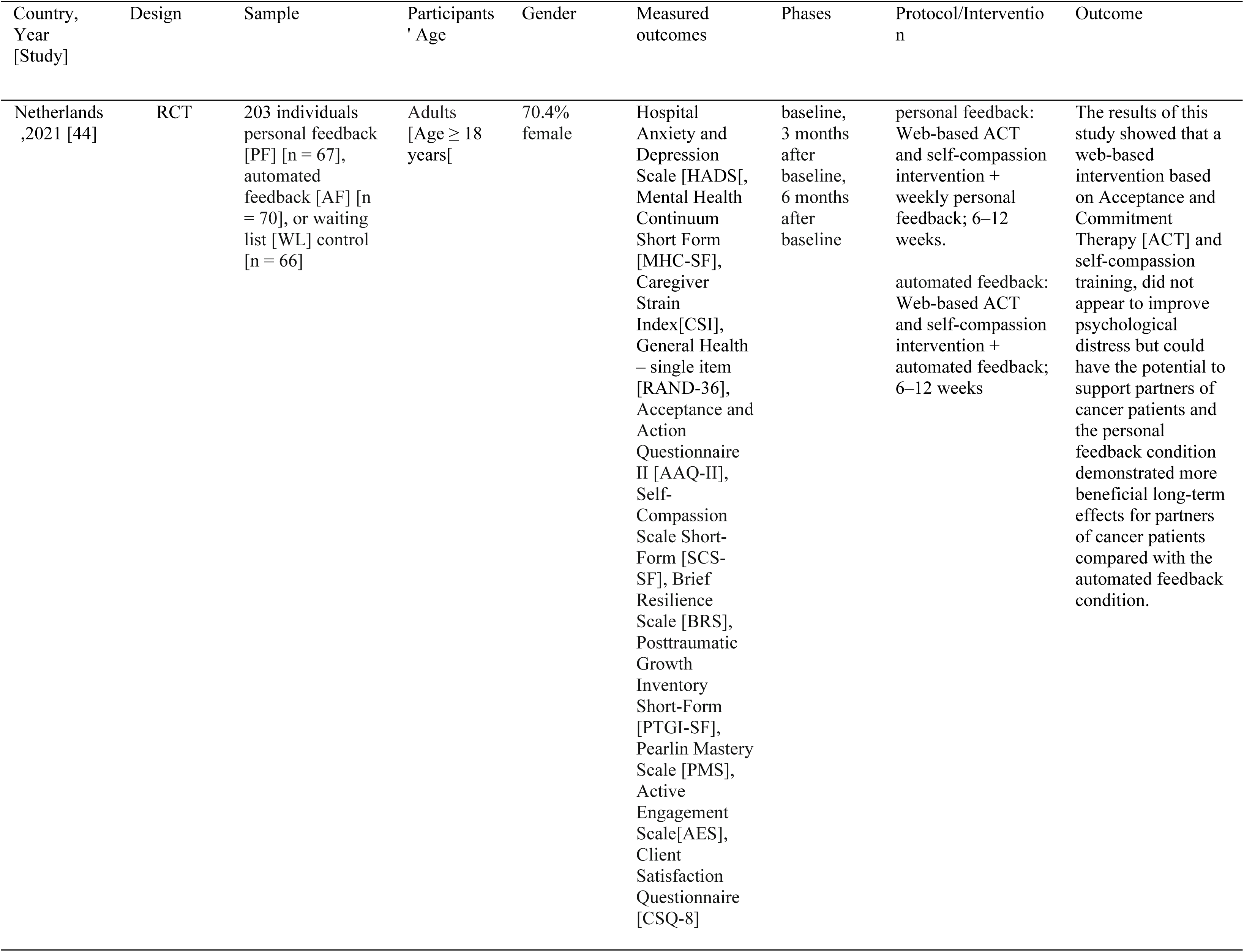
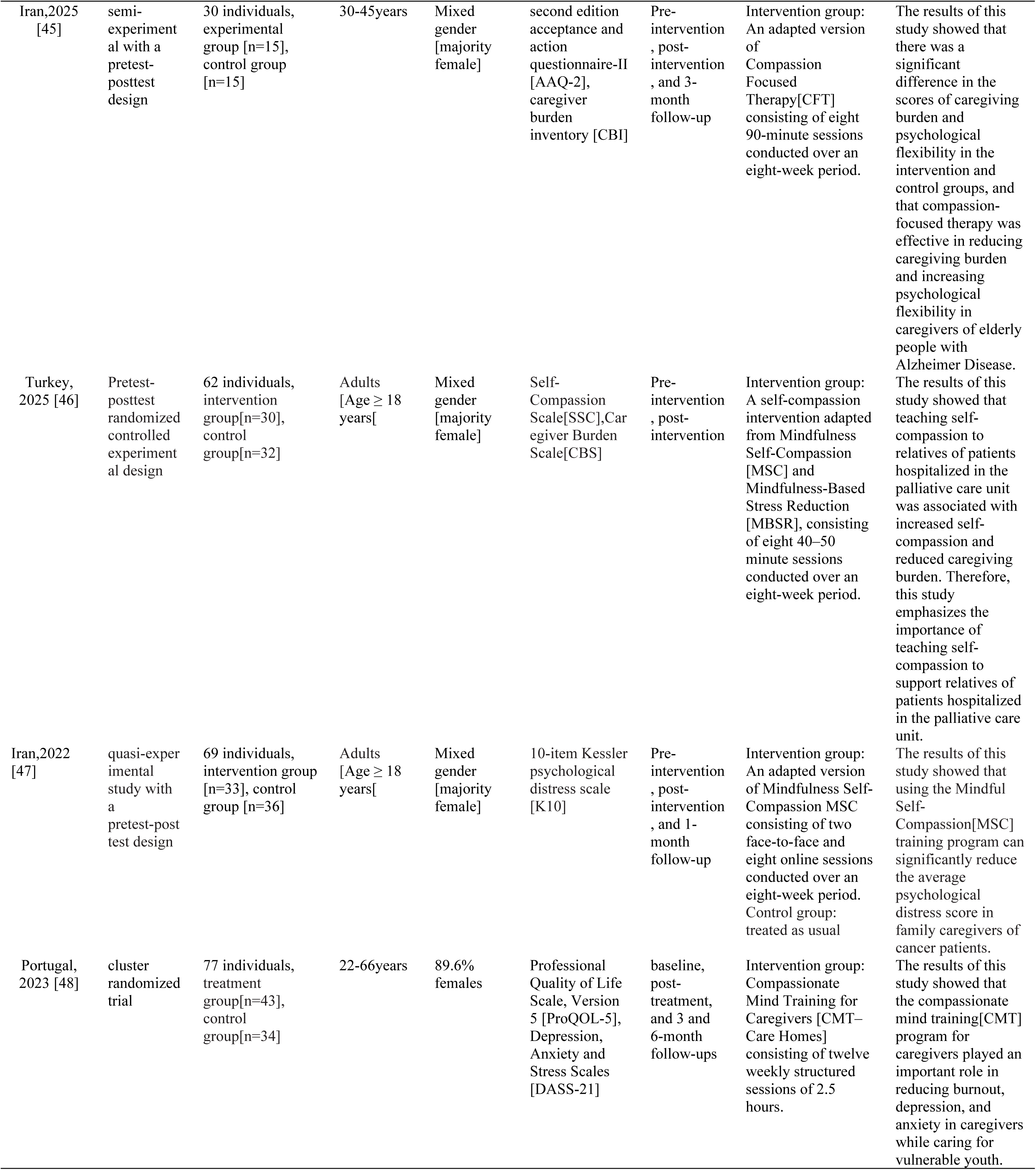
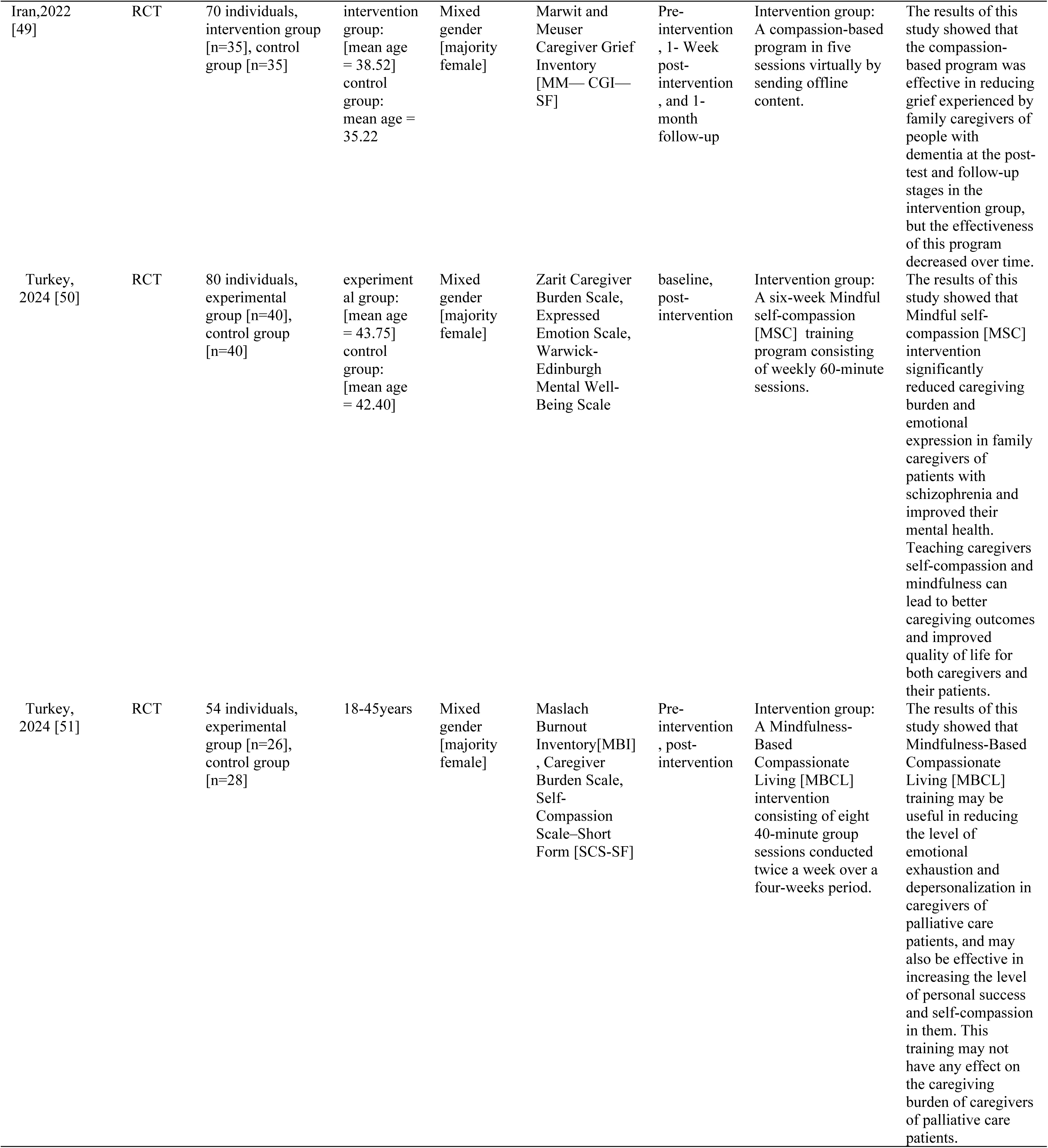
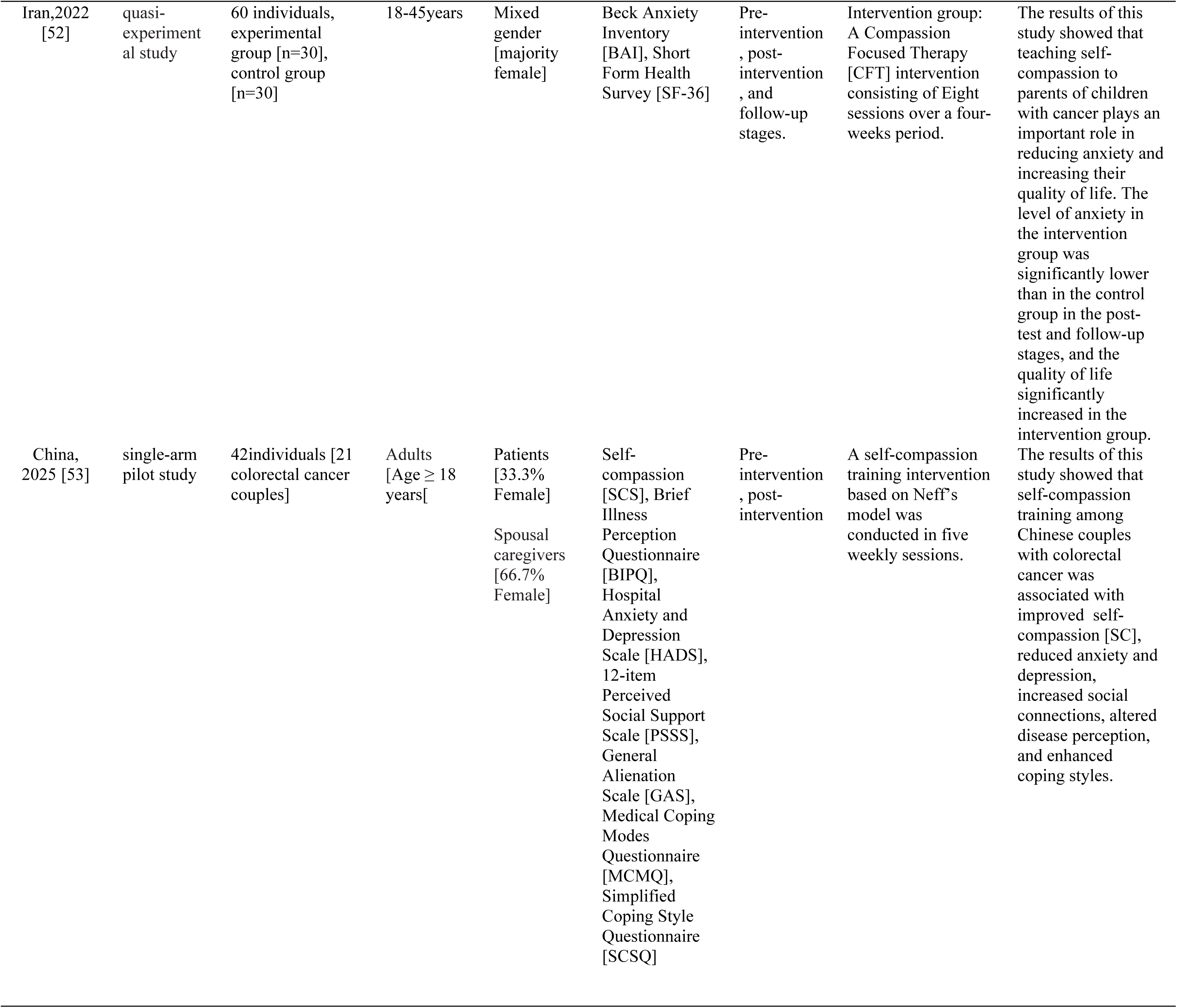
Summary of studies conducted using self-compassion training for caregivers or relatives of patients.

## Appendix 2

**Table 2.**
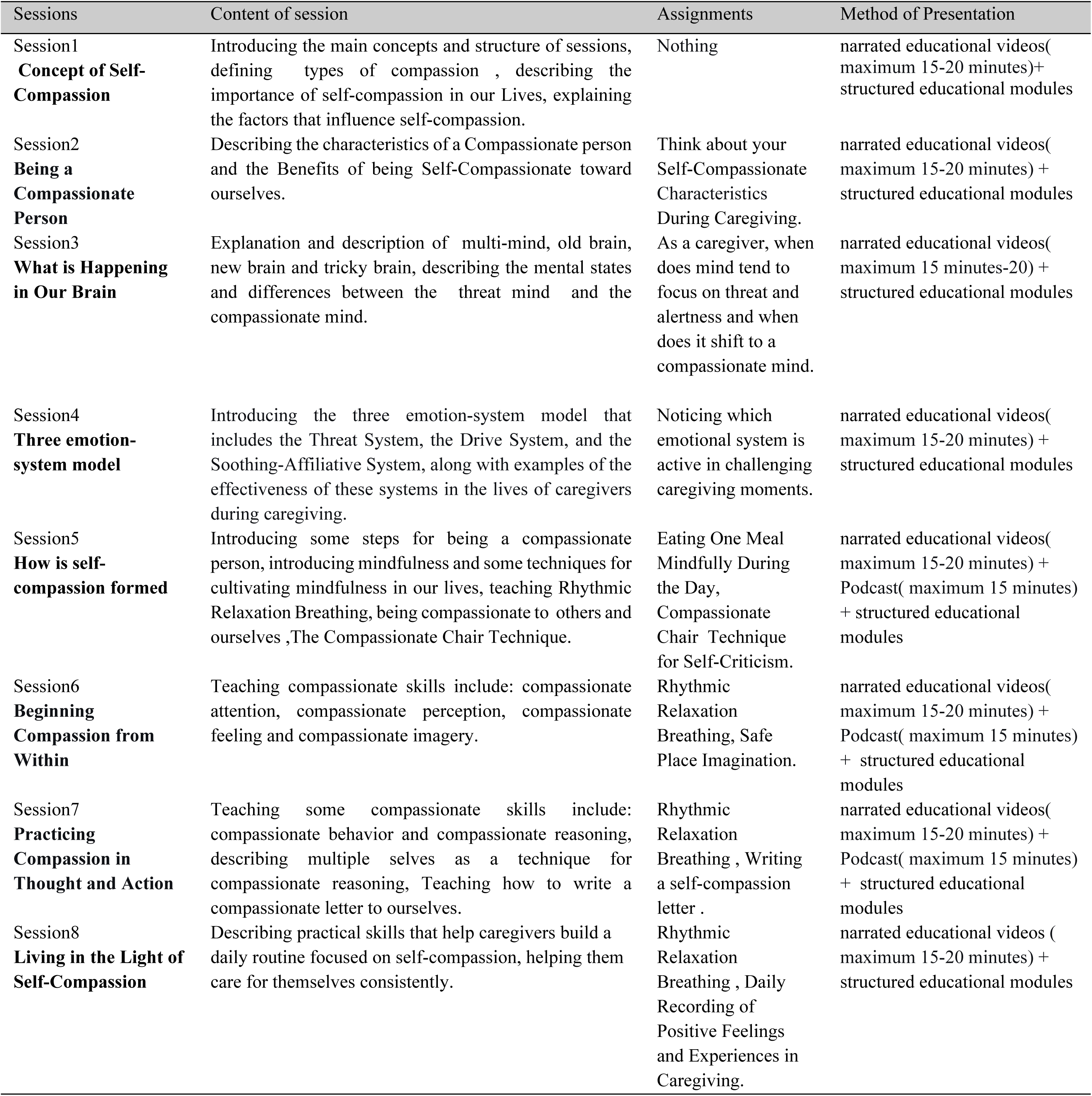
Sessions, Content of Sessions and Assignments of Compassion-Focused Therapy [Gillbert, 81].

## Appendix 3

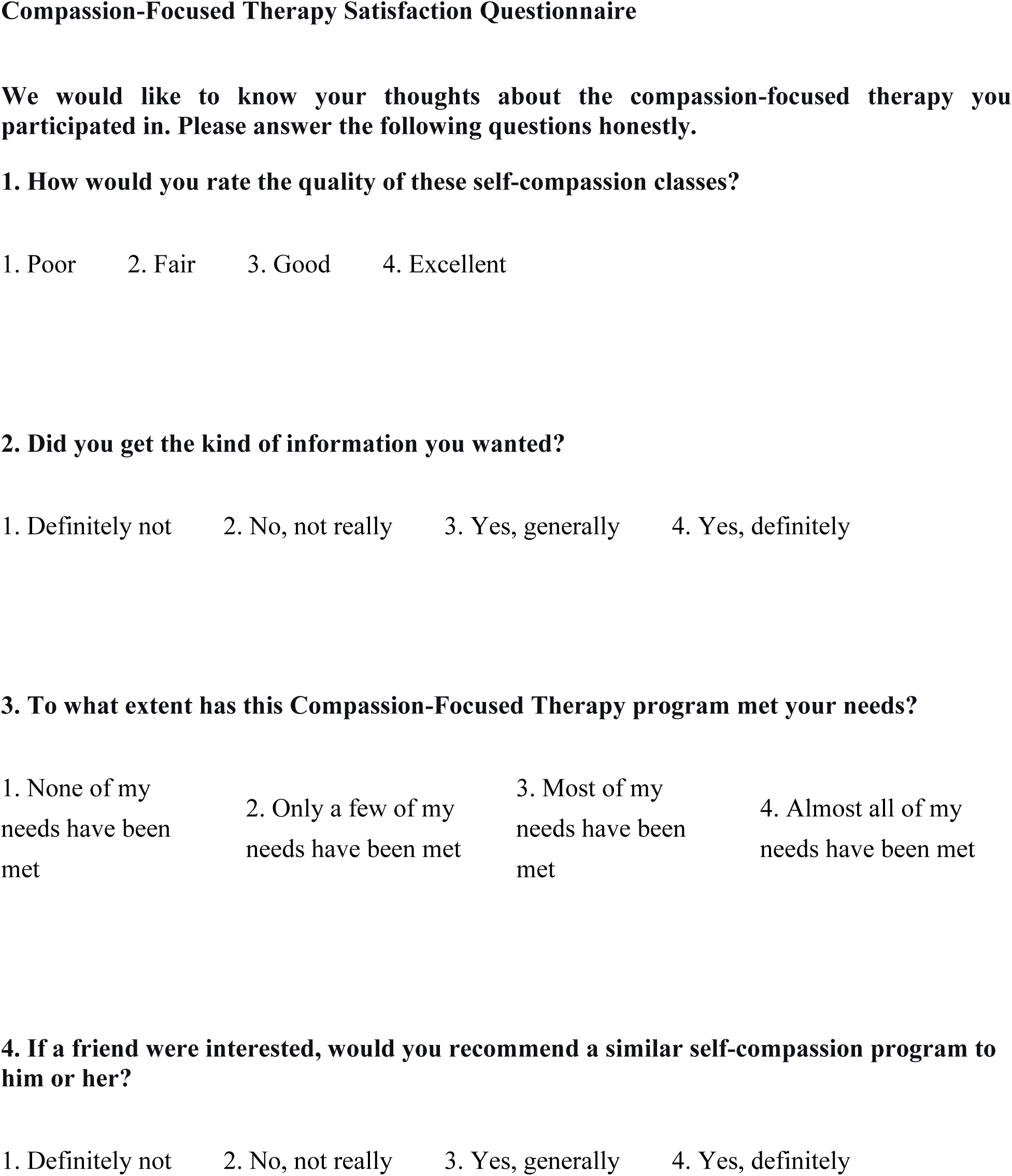
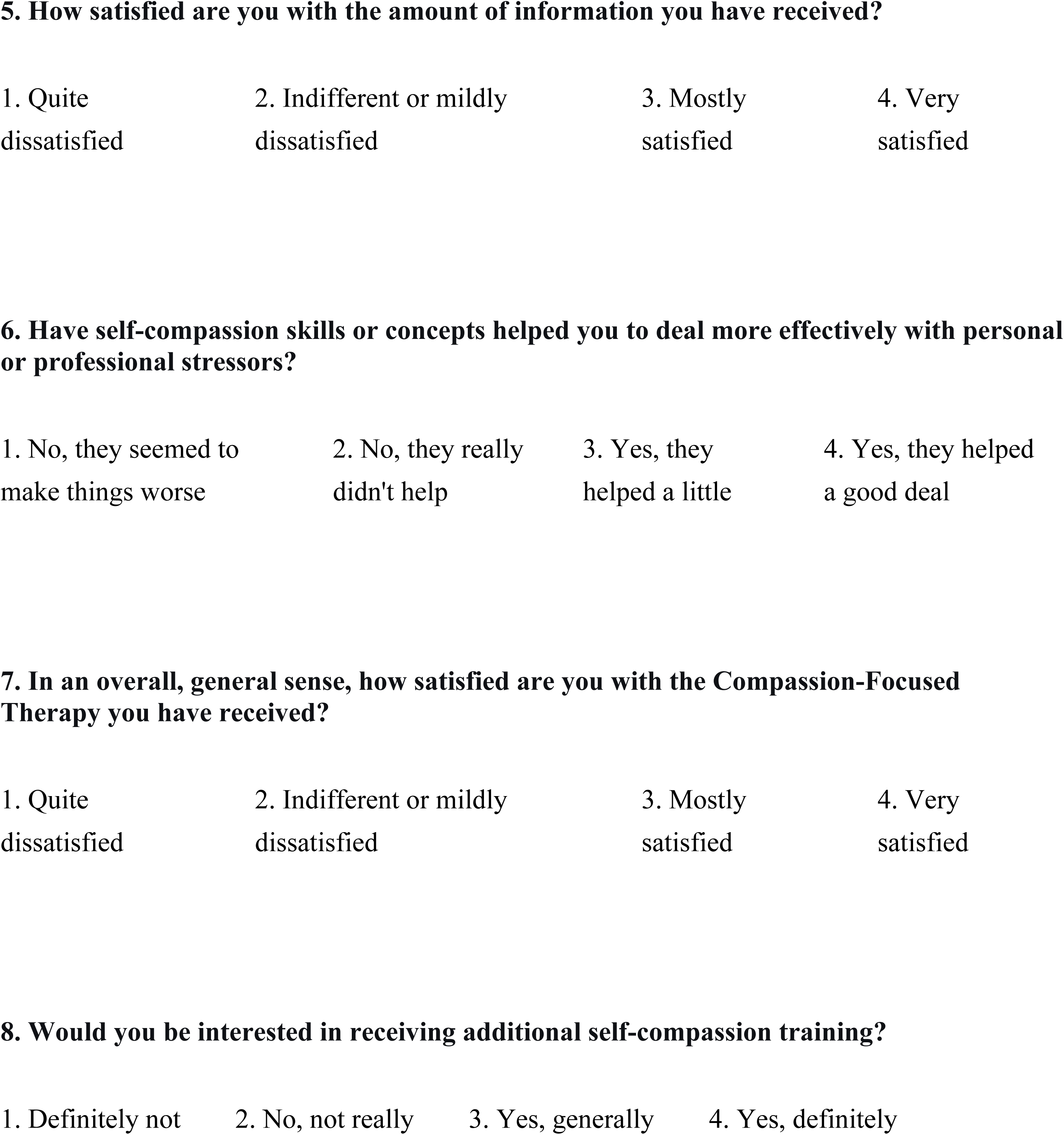

**3. To what extent has this Compassion-Focused Therapy program met your needs?**
|  |  |  |  |
| --- | --- | --- | --- |
| 1. None of my<br>needs have been<br>met | 2. Only a few of my<br>needs have been met | 3. Most of my<br>needs have been<br>met | 4. Almost all of my<br>needs have been met |
| 1. Quite<br>dissatisfied | 2. Indifferent or mildly<br>dissatisfied | 3. Mostly<br>satisfied | 4. Very<br>satisfied |
| 1. No, they seemed to<br>make things worse | 2. No, they really<br>didn't help | 3. Yes, they<br>helped a little | 4. Yes, they helped<br>a good deal |
| 1. Quite<br>dissatisfied | 2. Indifferent or mildly<br>dissatisfied | 3. Mostly<br>satisfied | 4. Very<br>satisfied |
| 1. Definitely not | 2. No, not really | 3. Yes, generally | 4. Yes, definitely |

## Appendix 4

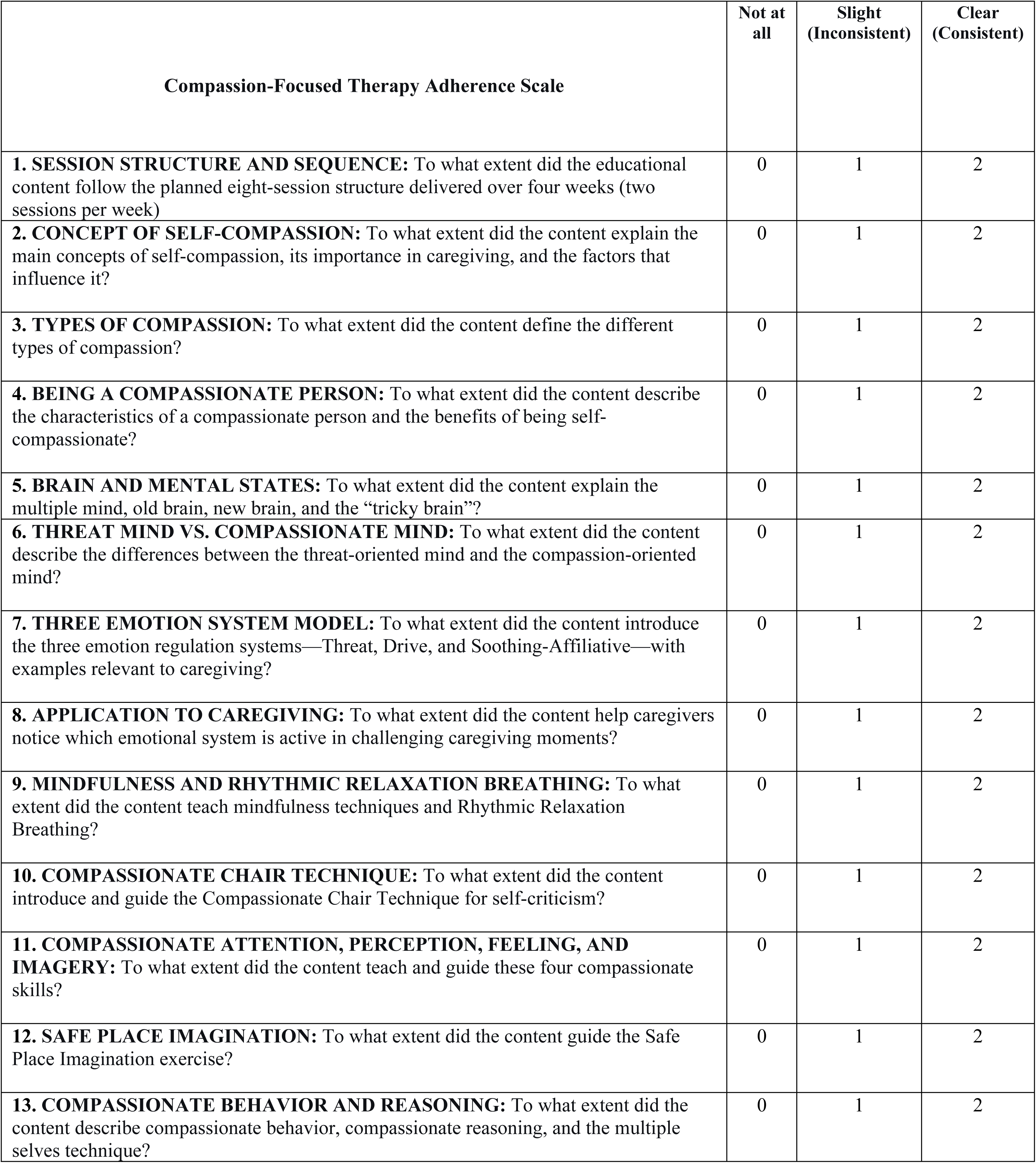
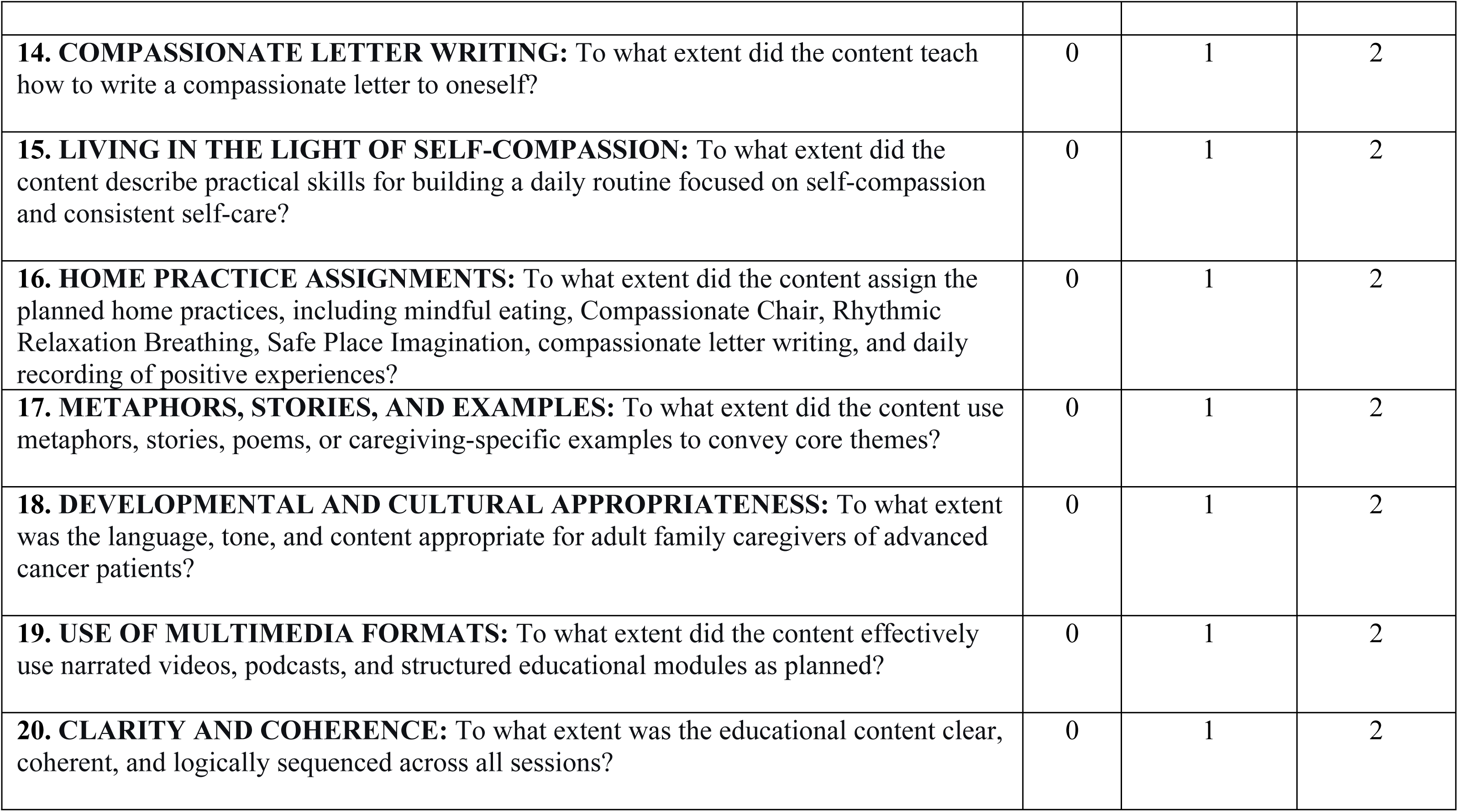

Web-based Self-Compassion Training for Cancer Caregivers Appendix 4
| Compassion-Focused Therapy Adherence Scale | Not at all | Slight (Inconsistent) | Clear (Consistent) |
| --- | --- | --- | --- |
| **1. SESSION STRUCTURE AND SEQUENCE:** To what extent did the educational content follow the planned eight-session structure delivered over four weeks (two sessions per week) | 0 | 1 | 2 |
| **2. CONCEPT OF SELF-COMPASSION:** To what extent did the content explain the main concepts of self-compassion, its importance in caregiving, and the factors that influence it? | 0 | 1 | 2 |
| **3. TYPES OF COMPASSION:** To what extent did the content define the different types of compassion? | 0 | 1 | 2 |
| **4. BEING A COMPASSIONATE PERSON:** To what extent did the content describe the characteristics of a compassionate person and the benefits of being self-compassionate? | 0 | 1 | 2 |
| **5. BRAIN AND MENTAL STATES:** To what extent did the content explain the multiple mind, old brain, new brain, and the “tricky brain”? | 0 | 1 | 2 |
| **6. THREAT MIND VS. COMPASSIONATE MIND:** To what extent did the content describe the differences between the threat-oriented mind and the compassion-oriented mind? | 0 | 1 | 2 |
| **7. THREE EMOTION SYSTEM MODEL:** To what extent did the content introduce the three emotion regulation systems—Threat, Drive, and Soothing-Affiliative—with examples relevant to caregiving? | 0 | 1 | 2 |
| **8. APPLICATION TO CAREGIVING:** To what extent did the content help caregivers notice which emotional system is active in challenging caregiving moments? | 0 | 1 | 2 |
| **9. MINDFULNESS AND RHYTHMIC RELAXATION BREATHING:** To what extent did the content teach mindfulness techniques and Rhythmic Relaxation Breathing? | 0 | 1 | 2 |
| **10. COMPASSIONATE CHAIR TECHNIQUE:** To what extent did the content introduce and guide the Compassionate Chair Technique for self-criticism? | 0 | 1 | 2 |
| **11. COMPASSIONATE ATTENTION, PERCEPTION, FEELING, AND IMAGERY:** To what extent did the content teach and guide these four compassionate skills? | 0 | 1 | 2 |
| **12. SAFE PLACE IMAGINATION:** To what extent did the content guide the Safe Place Imagination exercise? | 0 | 1 | 2 |
| **13. COMPASSIONATE BEHAVIOR AND REASONING:** To what extent did the content describe compassionate behavior, compassionate reasoning, and the multiple selves technique? | 0 | 1 | 2 |
| **14. COMPASSIONATE LETTER WRITING:** To what extent did the content teach how to write a compassionate letter to oneself? | 0 | 1 | 2 |
| **15. LIVING IN THE LIGHT OF SELF-COMPASSION:** To what extent did the content describe practical skills for building a daily routine focused on self-compassion and consistent self-care? | 0 | 1 | 2 |
| **16. HOME PRACTICE ASSIGNMENTS:** To what extent did the content assign the planned home practices, including mindful eating, Compassionate Chair, Rhythmic Relaxation Breathing, Safe Place Imagination, compassionate letter writing, and daily recording of positive experiences? | 0 | 1 | 2 |
| **17. METAPHORS, STORIES, AND EXAMPLES:** To what extent did the content use metaphors, stories, poems, or caregiving-specific examples to convey core themes? | 0 | 1 | 2 |
| **18. DEVELOPMENTAL AND CULTURAL APPROPRIATENESS:** To what extent was the language, tone, and content appropriate for adult family caregivers of advanced cancer patients? | 0 | 1 | 2 |
| **19. USE OF MULTIMEDIA FORMATS:** To what extent did the content effectively use narrated videos, podcasts, and structured educational modules as planned? | 0 | 1 | 2 |
| **20. CLARITY AND COHERENCE:** To what extent was the educational content clear, coherent, and logically sequenced across all sessions? | 0 | 1 | 2 |

## References

1. American Cancer Society. Global Cancer Facts & Figures. 5th ed. Atlanta: American Cancer Society; 2024. Available from: https://www.cancer.org/content/dam/cancer-org/research/cancer-facts-and-statistics/global-cancer-facts-and-figures/global-cancer-facts-and-figures-2024.pdf

2. Alizadeh Z, Rohani C, Rassouli M, Ilkhani M, Hazrati M. Bridging the gap and developing a home-based palliative care model for cancer patients. Asia-Pacific Journal of Oncology Nursing. 2024;11(11):100583. 10.1016/j.apjon.2024.100583 PMID: 39655227

3. International Agency for Research on Cancer. Global Cancer Observatory: Cancer Tomorrow. Estimated number of new cases from 2022 to 2045, both sexes, age [0–85+]: International Agency for Research on Cancer; 2022. Available from: https://gco.iarc.who.int/tomorrow/en/

4. International Agency for Research on Cancer. GLOBOCAN 2024: Iran, Islamic Republic of Fact Sheet. Lyon: International Agency for Research on Cancer; 2026. Available from: https://gco.iarc.who.int/media/globocan/factsheets/populations/364-iran-islamic-republic-of-fact-sheet.pdf

5. Barati H, Pourhoseingholi MA, Roshandel G, Nazari SSH. Bayesian age-period-cohort projection of cancers in Iran: a modeling study. BMC Cancer. 2024;24(1):1538. 10.1186/s12885-024-13289-0 PMID: 39696020

6. Goyanka R. Economic and non-economic burden of cancer: a propensity score matched analysis using household health survey data of India. Cancer Research, Statistics, and Treatment. 2021;4(1):29–36. 10.4103/crst.crst_6_21

7. Lewandowska A, Rudzki G, Lewandowski T, Rudzki S. The problems and needs of patients diagnosed with cancer and their caregivers. International journal of environmental research and public health. 2021;18(1):87. 10.3390/ijerph18010087 PMID: 33374440

8. Mirhosseini S, Imani Parsa F, Moghadam-Roshtkhar H, Basirinezhad MH, Ameri M, Ebrahimi H. Support based on psychoeducation intervention to address quality of life and care burden among caregivers of patients with cancer: a randomized controlled trial. Frontiers in Psychology. 2025;16:1430371. 10.3389/fpsyg.2025.1430371 PMID: 40083754

9. Odom JN, Henderson NL, Padalkar T, Ortiz Olguin E, Rocque GB. Roles performed by family caregivers to help individuals with cancer manage symptoms: A qualitative study. American Society of Clinical Oncology; 2025. 10.1200/OP.2025.21.10_suppl.302

10. Joolaee S, Ho A, Serota K, Hubert M, Buchman DZ. Medical assistance in dying legislation: Hospice palliative care providers’ perspectives. Nursing Ethics. 2022;29(1):231–44. 10.1177/09697330211012049 PMID: 34538192

11. Nates JL, Price KJ. Oncologic critical care. Springer; 2020. 10.1007/978-3-319-74588-6

12. Egici M, Can M, Toprak D, Öztürk G, Esen E, Özen B, et al. Burden and Burnout of Caregivers Whose Patients Treated at the Palliative Care Center. Journal of Academic Research In Nursing. 2019. 10.5222/jaren.2019.38247

13. Howard AF, Lynch K, Thorne S, Porcino A, Lambert L, De Vera MA, et al. Occupational and financial setbacks in caregivers of people with colorectal cancer: considerations for caregiver-reported outcomes. Current Oncology. 2022;29(11):8180–96. 10.3390/curroncol29110646 PMID: 36354706

14. Rezaei M, Keyvanloo Shahrestanaki S, Mohammadzadeh R, Aghili MS, Rajabi M, Abbasi M, et al. Caregiving consequences in cancer family caregivers: a narrative review of qualitative studies. Frontiers in Public Health. 2024; 12:1334842. 10.3389/fpubh.2024.1334842 PMID: 38584929

15. Kaddu-Mulindwa D, Jacob F, Schalbar C, Chrystofyllakis K, Bewarder M, Cloos J-M, et al. Quality of life and burden in family caregivers of advanced cancer patients in the Grand Duchy of Luxembourg: results from a single-centre study. Supportive Care in Cancer. 2026;34(4):296. 10.1007/s00520-026-10494-4 PMID: 41796255

16. Ringborg CH, Wengström Y, Schandl A, Lagergren P. The long-term experience of being a family caregiver of patients surgically treated for oesophageal cancer. Journal of Advanced Nursing. 2023;79(6):2259–68. 10.1111/jan.15580 PMID: 36779443

17. Hamedani B, Alavi M, Taleghani F, Fereidoonimoghadam M. Challenges of Help-Seeking in Iranian Family Caregivers of Patients with Cancer: A Qualitative Study. International Journal of Cancer Management. 2022;15(5):1. 10.5812/ijcm-127060

18. Washington KT, Craig KW, Parker Oliver D, Ruggeri JS, Brunk SR, Goldstein AK, et al. Family caregivers’ perspectives on communication with cancer care providers. Journal of psychosocial oncology. 2019;37(6):777–90. 10.1080/07347332.2019.1624674 PMID: 31204604

19. Papadakos J, Ugas M, Quartey NK, Papadakos C, Giuliani ME. Assessing the comprehensive training needs of informal caregivers of cancer patients: a qualitative study. Current Oncology. 2023;30(4):3845–58. 10.3390/curroncol30040291 PMID: 37185404

20. Biney A, Sarfo JO, Poku CA, Deegbe DA, Atibila F, Ti-enkawol Nachinab G, et al. Challenges and coping strategies when caring for terminally ill persons with cancer: perspectives of family caregivers. BMC Palliative Care. 2024;23(1):175. 10.1186/s12904-024-01518-z PMID: 39020352

21. Ma H, Zhao T, Ma Y, Yuen JWM, Ka Yan H, Yung JY, et al. Family caregivers’ lived experience of caring for hospitalised patients with cancer during the COVID-19 lockdown: A descriptive phenomenological study. Journal of clinical nursing. 2023;32(19-20):7509–18. 10.1111/jocn.16817 PMID: 37370254

22. Lim J, Cho H, Bunds KS, Lee C-W. Cancer family caregivers’ quality of life and the meaning of leisure. Health care for women international. 2021;42(7-9):1144–64. 10.1080/07399332.2020.1752214 PMID: 32490741

23. Hassankhani H, Eghtedar S, Rahmani A, Ebrahimi H, Whitehead B. A qualitative study on cancer care burden: experiences of Iranian family caregivers. Holistic nursing practice. 2019;33(1):17–26. 10.1097/HNP.0000000000000309 PMID: 30422921

24. Sigurlaugardottir SS, Sigurdardottir SH, Aspelund T, Bjornsdottir K, Jegermalm M, Olafsson K, et al. Elements of burden among informal caregivers of community-dwelling older adults receiving home care nursing: a cross-sectional study on health status, well-being, and gender differences. BMC geriatrics. 2025;25(1):623. 10.1186/s12877-025-06320-x PMID: 40813637

25. Oechsle K. Current advances in palliative & hospice care: Problems and needs of relatives and family caregivers during palliative and hospice care—An overview of current literature. Medical Sciences. 2019;7(3):43. 10.3390/medsci7030043 PMID: 30871105

26. Liu Z, Heffernan C, Tan J. Caregiver burden: A concept analysis. International journal of nursing sciences. 2020;7(4):438–45. 10.1016/j.ijnss.2020.07.012 PMID: 33195757

27. Su J-A, Chang C-C. Association between family caregiver burden and affiliate stigma in the families of people with dementia. International journal of environmental research and public health. 2020;17(8):2772. 10.3390/ijerph17082772 PMID: 32316454

28. Wong LP, Xu X, Alias H, Ting CY, Tan HM, Lin Y, editors. Patient-and caregiver-related factors affecting family caregiver burden of urologic cancer patients. Urologic Oncology: Seminars and Original Investigations; 2024: Elsevier. 10.1016/j.urolonc.2024.04.004 PMID: 38670816

29. Semere W, Althouse AD, Rosland A-M, White D, Arnold R, Chu E, et al. Poor patient health is associated with higher caregiver burden for older adults with advanced cancer. Journal of geriatric oncology. 2021;12(5):771–8. 10.1016/j.jgo.2021.01.002 PMID: 33478890

30. Pop RS, Mosoiu DV, Puia A, Tint D. Comparison of the burden evolution of the family caregivers for patients with cancer and nononcological diseases who need palliative care: A prospective longitudinal study. Palliative Medicine Reports. 2023;4(1): pmr. 2022.0067. 10.1089/pmr.2022.0067 PMID: 37483880

31. Au TH, Willis C, Reblin M, Peters KB, Nghiemphu PL, Taylor JW, et al. Caregiver burden by treatment and clinical characteristics of patients with glioblastoma. Supportive Care in Cancer. 2022;30(2):1365–75. 10.1007/s00520-021-06514-0 PMID: 34510238

32. Xia W, Liu T, Wu M, Luo Z, Guo D, Tian L. The trajectory of caregiver burden and the predictive role of sense of coherence among primary caregivers of colorectal cancer patients: a longitudinal study. BMC nursing. 2025;24(1):1202. 10.1186/s12912-025-03807-1 PMID: 41013519

33. Cui P, Yang M, Hu H, Cheng C, Chen X, Shi J, et al. The impact of caregiver burden on quality of life in family caregivers of patients with advanced cancer: a moderated mediation analysis of the role of psychological distress and family resilience. BMC Public Health. 2024;24(1):817. 10.1186/s12889-024-18321-3 PMID: 38491454

34. Abbasi A, Mirhosseini S, Basirinezhad MH, Ebrahimi H. Relationship between caring burden and quality of life in caregivers of cancer patients in Iran. Supportive Care in Cancer. 2020;28(9):4123–9. 10.1007/s00520-019-05240-y PMID: 31872293

35. Kim Y, Given BA. Quality of life of family caregivers of cancer survivors: across the trajectory of the illness. Cancer. 2008;112(S11):2556–68. 10.1002/cncr.23449 PMID: 18428199

36. Krug K, Miksch A, Peters-Klimm F, Engeser P, Szecsenyi J. Correlation between patient quality of life in palliative care and burden of their family caregivers: a prospective observational cohort study. BMC palliative care. 2016;15(1):4. 10.1186/s12904-016-0082-y PMID: 26767785

37. O’Hara RE, Hull JG, Lyons KD, Bakitas M, Hegel MT, Li Z, et al. Impact on caregiver burden of a patient-focused palliative care intervention for patients with advanced cancer. Palliative & supportive care. 2010;8(4):395–404. 10.1017/S1478951510000258 PMID: 20875202

38. Haroen H, Juniarti N, Sari CWM, Prista Sari S, Arovah NI, Pardosi JF, et al. Factors associated with caregiver burden in families of patients with palliative and chronic illness: a cross-sectional study. Journal of Multidisciplinary Healthcare. 2025:4497–510. 10.2147/JMDH.S533067 PMID: 40771865

39. Guo Y. Internet-delivered cognitive behavioral therapy versus standard information brochures in advanced cancer caregivers: randomized controlled trial. Internet Interventions. 2025:100891. 10.1016/j.invent.2025.100891 PMID: 41362329

40. Mosher CE, Secinti E, Wu W, Kashy DA, Kroenke K, Bricker JB, et al. Acceptance and commitment therapy for patient fatigue interference and caregiver burden in advanced gastrointestinal cancer: Results of a pilot randomized trial. Palliative medicine. 2022;36(7):1104–17. 10.1177/02692163221099610 PMID: 35637615

41. Faezeh TFT, Hakimeh AHA, Shahnaz NSN, Seyed MTSMT. Comparing the Effectiveness of Emotion-Focused Therapy and Compassion-Focused Therapy on the Belief in a Fair-Unfair World and Care Pressure in Caregivers of Cancer Patients. 2024.

42. Lloyd J, Muers J, Patterson TG, Marczak M. Self-compassion, coping strategies, and caregiver burden in caregivers of people with dementia. Clinical gerontologist. 2019;42(1):47–59. 10.1080/07317115.2018.1461162 PMID: 29723129

43. Neff KD, Faso DJ. Self-compassion and well-being in parents of children with autism. Mindfulness. 2015;6(4):938–47. 10.1007/s12671-014-0359-2

44. Köhle N, Drossaert CHC, Ten Klooster PM, Schreurs KMG, Hagedoorn M, Van Uden-Kraan CF, et al. Web-based self-help intervention for partners of cancer patients based on acceptance and commitment therapy and self-compassion training: a randomized controlled trial with automated versus personal feedback. Support Care Cancer. 2021;29(9):5115–25. 10.1007/s00520-021-06051-w PMID: 33608762

45. Madani SF, Farrokhzad P. The Effectiveness of Compassion-Focused Therapy on Caregiver Burden and Psychological Flexibility in Caregivers of Alzheimer’s Elderly. Iranian Journal of Ageing.0-. 10.32598/sija.2024.3786.1

46. Karakurt N, Erden Y, Turan GB, Bakir E. Invitation to Self-Compassion: Reshaping Burden of Care in the Light of Self-Compassion Training Given to Relatives of Patients Hospitalised in Palliative Care. Scandinavian Journal of Caring Sciences. 2025;39(2): e70038. 10.1111/scs.70038 PMID: 40411232

47. Noei FR, Atashi V, Ashouri E. The effect of an online mindfulness self-compassion training program on psychological distress in caregivers of patients with cancer. Nursing and Midwifery Studies. 2022;11(2):90–5. 10.4103/nms.nms_101_21

48. Santos L, Pinheiro MdR, Rijo D, editors. The effects of the compassionate mind training for caregivers on professional quality of life and mental health: outcomes from a cluster randomized trial in residential youth care settings. Child & Youth Care Forum; 2024: Springer. 10.1007/s10566-023-09749-6 PMID: 37360762

49. Jahani L, Abolhassani S, Babaee S, Omranifard V. Effects of a compassion-based program on the grief experienced by caregivers of people suffering from dementia: a randomized controlled clinical trial. BMC Nursing. 2022;21(1):198. 10.1186/s12912-022-00980-5 PMID: 35879751

50. Lök N, Bademli K. The Effect of a Mindful Self-Compassion Intervention on Burden, Express Emotion and Mental Well-Being in Family Caregivers of Patients with Schizophrenia: A Randomized Controlled Trial. Community Ment Health J. 2024;60(5):997-1005. 10.1007/s10597-024-01253-y PMID: 38393493

51. Kılıç D, Tosun Taşar P, Cengiz M. The effect of mindfulness-based compassionate living training for informal caregivers of palliative inpatients on burnout and caregiving burden: a randomized controlled trial. Health Education Research. 2024;39(4):313–22. 10.1093/her/cyae005 PMID: 38411949

52. Borjali M, Rahmani s, zakerzadeh P, Zeinali M. Effectiveness of Self-Compassion-Focused Therapy on Anxiety and Quality of Life of Parents of Children with Cancer. Clinical Psychology and Personality. 2022;19(2):25–32. 10.22070/cpap.2021.15184.1150

53. Yang X, Sun Z, Chen X, Qiu X, Mao J, Wang C, et al. Feasibility and acceptability of a couple-based self-compassion intervention for couples living with colorectal cancer: A pilot study. European Journal of Oncology Nursing. 2025:102899. 10.1016/j.ejon.2025.102899 PMID: 40466522

54. Neff KD. Self-compassion: Theory, method, research, and intervention. Annual review of psychology. 2023;74(1):193–218. 10.1146/annurev-psych-032420-031047 PMID: 35961039

55. Neff K. Self-compassion: An alternative conceptualization of a healthy attitude toward oneself. Self and identity. 2003;2(2):85–101. 10.1080/15298860309032

56. Neff KD, Germer CK. A pilot study and randomized controlled trial of the mindful self-compassion program. Journal of clinical psychology. 2013;69(1):28–44. 10.1002/jclp.21923 PMID: 23070875

57. Gilbert P. Introducing compassion-focused therapy. Advances in psychiatric treatment. 2009;15(3):199–208. 10.1192/apt.bp.107.005264

58. Gilbert P. The origins and nature of compassion focused therapy. British journal of clinical psychology. 2014;53(1):6–41. 10.1111/bjc.12043 PMID: 24588760

59. Wilson AC, Mackintosh K, Power K, Chan SW. Effectiveness of self-compassion related therapies: A systematic review and meta-analysis. Mindfulness. 2019;10(6):979–95. 10.1007/s12671-018-1037-6

60. Etemadi Shamsababdi P, Dehshiri GR. Self-compassion, anxiety and depression symptoms; the mediation of shame and guilt. Psychological reports. 2026;129(1):127–43. 10.1177/00332941241227525 PMID: 38251880

61. Hollis-Walker L, Colosimo K. Mindfulness, self-compassion, and happiness in non-meditators: A theoretical and empirical examination. Personality and Individual differences. 2011;50(2):222–7. 10.1016/j.paid.2010.09.033

62. Miller CL, Strachan SM. Understanding the role of mother guilt and self-compassion in health behaviors in mothers with young children. Women & health. 2020;60(7):763–75. 10.1080/03630242.2020.1713966 PMID: 31937201

63. Gilbert P, Procter S. Compassionate mind training for people with high shame and self-criticism: Overview and pilot study of a group therapy approach. Clinical Psychology & Psychotherapy: An International Journal of Theory & Practice. 2006;13(6):353–79. 10.1002/cpp.507

64. Mohamed Hussin NA, Mohd Sabri NS. A qualitative exploration of the dynamics of guilt experience in family cancer caregivers. Supportive Care in Cancer. 2023;31(11):659. 10.1007/s00520-023-08060-3 PMID: 37889352

65. Spillers RL, Wellisch DK, Kim Y, Matthews A, Baker F. Family caregivers and guilt in the context of cancer care. Psychosomatics. 2008;49(6):511–9. 10.1176/appi.psy.49.6.511 PMID: 19122128

66. Le Z, Yaofeng Z, Shuo L, Lisha T, Wenya H, Huihong Z, et al. The Mediating Role of Caregiver Guilt in the Relationship Between Stroke Patients’ Functional Status and Caregiver Burden: A Correlational Study. International Journal of Nursing Studies Advances. 2025:100437. 10.1016/j.ijnsa.2025.100437 PMID: 41234675

67. Valdez CE, Lilly MM. Modes of processing trauma: Self-compassion buffers affective guilt. Mindfulness. 2019;10(5):824–32. 10.1007/s12671-018-1035-8

68. Doerwald F, Stalling I, Recke C, Busse H, Shrestha R, Rach S, et al. A rapid review of digital approaches for the participatory development of health-related interventions. Frontiers in Public Health. 2024; 12:1461422. 10.3389/fpubh.2024.1461422 PMID: 39678234

69. Kim M, Tan KR, Coombs LA. Effects of web-based interventions on cancer caregivers’ burden and quality of life: A systematic review and meta-analysis. Palliative & Supportive Care. 2025;23: e134. 10.1017/S1478951525100370 PMID: 40676719

70. Sang E, Hirschman KB, Bowles KH. Feasibility, usability, and acceptability of psychoeducational videoconferencing interventions for informal caregivers: A systematic review of randomized controlled trials. Research in nursing & health. 2024 Feb;47(1):60–81. 10.1002/nur.22358 PMID: 38069607

71. Zhai S, Chu F, Tan M, Chi NC, Ward T, Yuwen W. Digital health interventions to support family caregivers: An updated systematic review. Digital health. 2023 May; 9:20552076231171967. 10.1177/20552076231171967 PMID: 37223775

72. Khosrobeigi M, Hafezi F, Naderi F, Ehteshamzadeh P. Effectiveness of self-compassion training on hopelessness and resilience in parents of children with cancer. Explore. 2022;18(3):357–61. 10.1016/j.explore.2021.04.003 PMID: 33906814

73. Tourani F, Aghaee H, Nouhi S, Tabatabaee SM. Comparing the Effectiveness of Emotion-Oriented Therapy and Compassion-Based Therapy on Premature Bereavement and Psychological Distress of Caregivers of Cancer Patients. Rooyesh-e-Ravanshenasi Journal (RRJ). 2024;13(7):131–40. Available from: http://frooyesh.ir/article-1-5568-en.html

74. Le KT, Emerson GA. Caregiver burden, coping strategies, and quality of life among family caregivers of adult cancer patients: A multiple regression and mediation analysis in Southern Vietnam. Tạp chí Khoa học Điều dưỡng. 2025;8(05):137–50. 10.54436/jns.2025.05.1098

75. Chan A-W, Boutron I, Hopewell S, Moher D, Schulz KF, Collins GS, et al. SPIRIT 2025 statement: updated guideline for protocols of randomised trials. The Lancet. 2025;405(10491): e19-e27. 10.1136/bmj-2024-081477 PMID: 40294953

76. Faul F, Erdfelder E, Lang A-G, Buchner A. G* Power 3: A flexible statistical power analysis program for the social, behavioral, and biomedical sciences. Behavior research methods. 2007;39(2):175–91. 10.3758/bf03193146 PMID: 17695343

77. Lim WP, Loo JY, Lee K, Pui HM, Ting TT. The impact of social media on student’s academic performance: A survey on TAR UC computing students in Malaysia during COVID-19 pandemic. InInternational Conference on Digital Transformation and Applications (ICDXA) 2021 Oct (Vol. 25, pp. 161-167).

78. Weeratunga E, Goonewardena S, Meegoda L. Comprehensive needs assessment tool for informal cancer caregivers (CNAT-ICs): Instrument development and cross-sectional validation study. International Journal of Nursing Studies Advances. 2024; 7:100240. 10.1016/j.ijnsa.2024.100240 PMID: 39391564

79. Finlay-Jones AL, Parkinson A, Sirois F, Perry Y, Boyes M, Rees CS. Web-based self-compassion training to improve the well-being of youth with chronic medical conditions: Randomized controlled trial. Journal of Medical Internet Research. 2023;25: e44016. 10.2196/44016 PMID: 37703081

80. Sahraian K, Abdollahpour Ranjbar H, Namavar Jahromi B, Cheung HN, Ciarrochi J, Habibi Asgarabad M. Effectiveness of mindful self-compassion therapy on psychopathology symptoms, psychological distress and life expectancy in infertile women treated with in vitro fertilization: a two-arm double-blind parallel randomized controlled trial. BMC psychiatry. 2024 Mar 1;24(1):174. 10.1186/s12888-023-05411-6 PMID: 38429659

81. Gilbert P. Compassion focused therapy: Distinctive features: Routledge; 2010. 10.4324/9780203851197

82. Neff KD. The development and validation of a scale to measure self-compassion. Self and identity. 2003;2(3):223–50. 10.1080/15298860309027

83. Azizi A, Mohammadkhani P, Lotfi S, Bahramkhani M. The validity and reliability of the Iranian version of the Self-Compassion Scale. Practice in Clinical Psychology. 2013;1(3):149–55. http://jpcp.uswr.ac.ir/article-1-83-en.html

84. Novak M, Guest C. Application of a multidimensional caregiver burden inventory. Gerontologist. 1989;29(6):798–803. 10.1093/geront/29.6.798 PMID: 2516000

85. Shafiezadeh A, Heravi-Karimooi M, Mirzaee A, Rejeh N, Sharif Nia H, Montazeri A. Psychometric characteristics of the Iranian Caregiver Burden Inventory (CBI) in caregivers of elderly patients with Alzheimer. Health Qual Life Outcomes. 2020;18(1):255. 10.1186/s12955-020-01509-7 PMID: 32727503

86. Weitzner M, Meyers C, Steinbruecker S, Saleeba A, Sandifer S. Developing a care giver quality-of-life instrument. Preliminary steps. Cancer practice. 1997;5(1):25–31. PMID: 9128493

87. Weitzner MA, Jacobsen PB, Wagner H, Jr., Friedland J, Cox C. The Caregiver Quality of Life Index-Cancer (CQOLC) scale: development and validation of an instrument to measure quality of life of the family caregiver of patients with cancer. Qual Life Res. 1999;8(1-2):55–63. 10.1023/a:1026407010614 PMID: 10457738

88. Khanjari S, Oskouie F, Langius-Eklöf A. Psychometric testing of the Caregiver Quality of Life Index-Cancer scale in an Iranian sample of family caregivers to newly diagnosed breast cancer women. J Clin Nurs. 2012;21(3-4):573–84. 10.1111/j.1365-2702.2011.03850.x PMID: 21923672

89. Attkisson CC, Zwick R. The client satisfaction questionnaire. Psychometric properties and correlations with service utilization and psychotherapy outcome. Eval Program Plann. 1982;5(3):233–7. 10.1016/0149-7189(82)90074-x PMID: 10259963

90. Matsubara C, Green J, Astorga LT, Daya EL, Jervoso HC, Gonzaga EM, et al. Reliability tests and validation tests of the client satisfaction questionnaire (CSQ-8) as an index of satisfaction with childbirth-related care among Filipino women. BMC pregnancy and childbirth. 2013 Dec 17; 13:235. doi: 10.1186/1471-2393-13-235 PMID: 24341288

91. Semple RJ, Sears R. Mindfulness-based cognitive therapy for children (MBCT)—adherence scale. Unpublished assessment scale. University of Southern California, Los Angeles. 2014.

92. Pp R. Human experimentation. Code of ethics of the world medical association. Declaration of Helsinki. British medical journal. 1964;2(5402):177-. 10.1136/bmj.2.5402.177 PMID: 14150898

93. Williams JR. The Declaration of Helsinki and public health. Bulletin of the World Health Organization. 2008;86:650–2. 10.2471/blt.08.050955 PMID: 18797627

94. Nysaeter TM, Olsson C, Sandsdalen T, Hov R, Larsson M. Family caregivers’ preferences for support when caring for a family member with cancer in late palliative phase who wish to die at home–a grounded theory study. BMC palliative care. 2024;23(1):15. 10.1186/s12904-024-01350-5 PMID: 38212707

95. Li Y, Li J, Zhang Y, Ding Y, Hu X. The effectiveness of e-Health interventions on caregiver burden, depression, and quality of life in informal caregivers of patients with cancer: A systematic review and meta-analysis of randomized controlled trials. International journal of nursing studies. 2022; 127:104179. 10.1016/j.ijnurstu.2022.104179 PMID: 35124473

96. Savitri W, Suwarno S. Burden on Family Caregivers Caring for Patients with Cancer in Indonesia. Proceedings Series on Health & Medical Sciences. 2021;2:77–81. 10.30595/pshms.v2i.226

97. Arian M, Masoumi R, Shojaei SF. Feelings of Shame and Guilt in Patients with Cancer and Their Family Caregivers, Letter. 2021. 10.5812/ijcm.117159

98. Ribeiro M, Carvalho PS, Torres A, Ferreira D. Guilt and Depressive Symptoms: The Mediating Role of Self-Compassion in Perinatal Loss. OMEGA-Journal of Death and Dying. 2026:00302228251415137. 10.1177/00302228251415137 PMID: 41514173

99. Wakelin KE, Perman G, Simonds LM. Effectiveness of self-compassion-related interventions for reducing self-criticism: A systematic review and meta-analysis. Clinical psychology & psychotherapy. 2022;29(1):1–25. 10.1002/cpp.2586 PMID: 33749936

100. Berg MN, Ngune I, Schofield P, Grech L, Juraskova I, Strasser M, et al. Effectiveness of online communication skills training for cancer and palliative care health professionals: a systematic review. Psycho-Oncology. 2021;30(9):1405–19. 10.1002/pon.5702 PMID: 33909328

